# AutoCumulus: an Automated Mammographic Density Measure Created Using Artificial Intelligence

**DOI:** 10.1101/2024.02.01.24302158

**Authors:** Osamah Al-qershi, Tuong L Nguyen, Michael S Elliott, Daniel F Schmidt, Enes Makalic, Shuai Li, Samantha K Fox, James G Dowty, Carlos A Peña-Solorzano, Chun Fung Kwok, Yuanhong Chen, Chong Wang, Jocelyn Lippey, Peter Brotchie, Gustavo Carneiro, Davis J McCarthy, Yeojin Jeong, Joohon Sung, Helen ML Frazer, John L Hopper

**Affiliations:** Centre for Epidemiology & Biostatistics, Melbourne School of Population and Global Health, University of Melbourne, Victoria, Australia; Bioinformatics and Cellular Genomics Unit, St Vincent’s Institute of Medical Research, Victoria, Australia; Melbourne Integrative Genomics, School of Mathematics and Statistics/School of BioSciences, Faculty of Science, University of Melbourne, Victoria, Australia; Faculty of Information Technology, Monash University, Clayton, Victoria, Australia; Centre for Cancer Genetic Epidemiology, Department of Public Health and Primary Care, University of Cambridge, Cambridge, UK; Precision Medicine, School of Clinical Sciences at Monash Health, Monash University, Clayton, Victoria, Australia; Murdoch Children’s Research Institute, Royal Children’s Hospital, Parkville, Victoria, Australia; School of Computer Science, Australian Institute for Machine Learning, University of Adelaide, South Australia, Australia; Department of Surgery, St Vincent’s Hospital Melbourne, Victoria, Australia; Faculty of Medicine, Dentistry and Health Sciences - University of Melbourne, Victoria, Australia; St Vincent’s BreastScreen, St Vincent’s Hospital Melbourne, Victoria, Australia; Department of Radiology, St Vincent’s Hospital Melbourne, Victoria, Australia; Centre for Vision, Speech and Signal Processing, University of Surrey, United Kingdom; Genome & Health Data Lab, Seoul National University School of Public Health, Seoul, Korea; BreastScreen Victoria, Victoria, Australia

## Abstract

**Background:** Mammographic (or breast) density is an established risk factor for breast cancer. There are a variety of approaches to measurement including quantitative, semi-automated and automated approaches. We present a new automated measure, AutoCumulus, learnt from applying deep learning to semi-automated measures.

**Methods:** We used mammograms of 9,057 population-screened women in the BRAIx study for which semi-automated measurements of mammographic density had been made by experienced readers using the CUMULUS software. The dataset was split into training, testing, and validation sets (80%, 10%, 10%, respectively). We applied a deep learning regression model (fine-tuned ConvNeXtSmall) to estimate percentage density and assessed performance by the correlation between estimated and measured percent density and a Bland-Altman plot. The automated measure was tested on an independent CSAW-CC dataset in which density had been measured using the LIBRA software, comparing measures for left and right breasts, sensitivity for high sensitivity, and areas under the receiver operating characteristic curve (AUCs).

**Results:** Based on the testing dataset, the correlation in percent density between the automated and human measures was 0.95, and the differences were only slightly larger for women with higher density. Based on the CSAW-CC dataset, AltoCumulus outperformed LIBRA in correlation between left and right breast (0.95 versus 0.79; P<0.001), specificity for 95% sensitivity (13% versus 10% (P<0.001)), and AUC (0.638 cf. 0.597; P<0.001).

**Conclusion:** We have created an automated measure of mammographic density that is accurate and gives superior performance on repeatability within a woman, and for prediction of interval cancers, than another well-established automated measure.

## 1. Introduction

Breast cancer is a prevalent and potentially life-threatening disease that affects millions of women worldwide. It is the leading cause of cancer-related deaths among women (Ferlay et al., 2020). However, timely detection and treatment can significantly improve outcomes. To address this, many developed countries have implemented large-scale mammography screening programs, advising women to begin screening between the ages of 40 and 50 (Arefan et al., 2020). Digital mammography is commonly now used for early detection, and assessing mammographic density is a crucial aspect of this process.

Mammographic (of breast) density is the defined in terms of the regions on a mammogram that are “white or bright”, as distinct from dark. It is presumed that the dense areas represent glandular and connective tissue rather than fatty tissue. Mammographic density is assessed only through mammographic imaging and is not related to the way the breasts feel.

Mammographic density poses challenges in breast cancer detection. Dense breast tissue appears white on mammograms, similar to cancerous tissue, making it more challenging to detect tumors during routine screening (Nazari & Mukherjee, 2018). This similarity increases the chances of false negatives, where cancers are missed in mammograms. Consequently, women with dense breasts might benefit from additional imaging tests, such as ultrasound or magnetic resonance imaging (MRI), to ensure early detection and accurate diagnosis (Grin et al., 2009). In this regard, fortunately mammographic density decreases with increasing age and with increasing body size (such as body mass index (BMI) or breast size), making screening more effective for older women.

Against this, many studies have found that mammographic density predicts future risk of breast cancer for screening-aged women. This appears to be paradoxical given that age and BMI are both associated with increasing breast risk for this specific population. Nevertheless, these associations exist even after adjusting for age and BMI, irrespective of how density is measured (D’Orsi et al., 2018; Engmann et al., 2017; McCormack & dos Santos Silva, 2006) (Lu et al., 2022)). Either way, the risk association is stronger for ‘interval’ cancers diagnosed in the next two or three years (Nguyen et al., Breast Cancer Res 2018)), thought to be at least in part due the role of masking existing cancers mentioned above.

There are a variety of approaches to measurement including quantitative, semi-automated and automated approaches. One qualitative approach is the BI-RADS density categorisation in which radiologists assign mammographic density into four groups referred to as: fatty (A), scattered fibroglandular (B), heterogeneously dense (C), or extremely dense (D). Wording to include visual assessments of percent density (percentage of the breast area covered by dense regions) in this process has been recently dropped.

Boyd and Yaffe developed a semi-automated way to segment regions and measure the area covered by what are considered dense regions and the total breast area using the computer program CUMULUS ((Boyd et al., 2007)). The human operator uses a toggle to delineate what they consider are the dense regions, leading to a measure of dense area. Percent mammographic density is then defined as the dense area / total breast area as a percentage.

More recently, automated methods for measuring mammographic density have been developed using what might be considered as artificial intelligence (AI) approaches. For example, the freely available Laboratory for Individualized Breast Biodensity Assessment (LIBRA) software package (Keller et al., 2015) used an adaptive multi-class fuzzy c-means algorithm to identify and partition the mammographic breast tissue area into multiple regions of similar intensity which were then aggregated by a support-vector machine classifier to obtain a measure of percent density. This has been found to have correlations with measures using the CUMULUS software of 0.77–0.84, and of 0.85–0.90 with measures using VOLPARA, a commercially available software based on a physics-based model (Gastounioti et al., 2020).

Kallenberg and colleagues used a convolutional sparse autoencoder to learn features from manually segmented dense areas created by a radiologist (Kallenberg et al., 2016). The correlation in percent density between their algorithm and the manual measure was 0.85. It is noteworthy that the algorithm’s segmentation performance was deemed suboptimal given the Dice coefficient, averaged across images, was only 0.63.

Lee and Nishikawa used the VGG16 network to segment the breast and the dense fibro glandular areas (Lee & Nishikawa, 2018). The correlation of their measures with those using LIBRA were in the range of 0.6 to 0.7. Other approaches to measuring density used applied conditional Generative Adversarial Networks (cGAN) (Saffari et al., 2020) and simple pulse coupled neural network (SPCNN) (Qi et al., 2021).

In this paper we devised AutoCumulus, a novel, fully automated approach for estimating mammographic density, as measured using CUMULUS, by applying deep learning to a large dataset of mammograms and CUMULUS measures. We employed a transfer-learning technique—fine-tuning—to train our algorithm. We then compared the percent density estimates of our model with those generated by human assessments and evaluated performance against the publicly available algorithm LIBRA algorithm using a large independent dataset.

## 2. Methods

### 2.1 Dataset

The dataset consists of 9,057 mammograms selected from the BRAIx program’s ADMANI dataset (Frazer et al., 2022) which contains over 4 million mammograms for 630,000 women. This population-based collection supports AI development for improved breast cancer detection and risk-based screening in Australia. The selected mammograms are from 6,781 women, comprising 1,381 women with and 5,400 without breast cancer. All selected mammograms were craniocaudal (CC) views to avoid the problem of the pectoral muscle in the mediolateral oblique (MLO) views. All images were generated using machines of the same manufacturer.

### 2.2 Semi-Automatic Mammographic Density Measures

Mammographic density had been measured on all images using the CUMULUS software by experienced measurers with high repeatability as in previous studies (REFs to Nguyen et al. papers). By moving a toggle to create a threshold that defines the outer limits of the breast image or a pixel brightness, the CUMULUS software automatically identifies the areas above and below that threshold by drawing a contour line; for example, the green line in Figure 1 outlines the dense regions.

**Figure 1.**
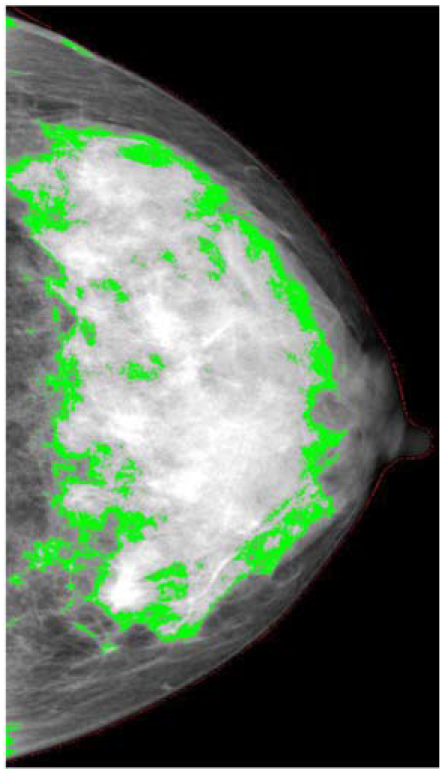
An example of a measurement of dense area (the area with the green edges) using the CUMULUS software giving a percent mammographic density of 58% (Nguyen et al., 2015).

To construct and assess the AI-trained model, we partitioned a total of 9,057 images, assigning 7,278, 888, and 891 mammograms exclusively to training, testing, and validation datasets, representing 80%, 10%, and 10% of the complete dataset, respectively. The three subsets were created randomly while maintaining the same proportion of age groups and affected-to-unaffected ratio in each dataset.

An experienced single observer, who was blinded to all identifying details, conducted the measurement of mammographic density using the computer-assisted thresholding technique, Cumulus (Imaging Research Program, Sunnybrook Health Sciences Centre, University of Toronto, Toronto, Canada). Cumulus provides direct measurements of the total breast area and the area occupied by mammographically dense tissue, as assessed by the observer. The non-dense area is subsequently calculated by subtracting the dense area from the total breast area. The percentage density is derived by dividing the dense area by the total breast area. Notably, this method, as demonstrated in previous studies (Byng et al., 1998), has exhibited reliability and high reproducibility.

### 2.3 Deep Learning Regression Model

We used a deep learning regression model to create an automated estimate of percent density from the semi-automated measures created using the CUMULUS software by using the pre-trained ConvNeXtSmall (Liu et al., 2022) network as the backbone of our model and fine-tuning. After convolutional layers, a Global Average Pooling layer was applied, followed by a densely connected layer (32 units) with ReLU activation and L1/L2 regularization for capturing intricate features. The output layer, consisting of a single neuron, is constrained to non-negativity. This approach learns hierarchical features from images while mitigating overfitting through regularization techniques and leveraging pre-trained knowledge from ImageNet.

### 2.4 Training Setup

The mammographic images typically had dimensions of approximately 3000 × 4000 pixels and were of 16-bit depth, which would necessitate substantial computational resources for model training. To address such computational demands and dimensionality challenges, we employed a preprocessing step in which all images were resized to a standardized dimension of 1024 × 768 pixels while maintaining the original aspect ratio. Additionally, the pixel intensity values were normalized to the [0,1] range, facilitating more efficient model training while preserving the essential information encapsulated in the images.

The proposed model was trained using the Adaptive Moment Estimation (Adam) optimizer, employing a learning rate of 0.0001, a decay rate of 0.90, and decay steps at 200 intervals. The training process utilized a batch size of 8, incorporating image augmentation techniques such as random horizontal and vertical flipping, along with a 5% brightness adjustment. Binary cross-entropy was employed as the loss function, and the training process was iterated for a maximum of 30 epochs to fine-tune the model’s parameters.

The model was constructed using Python 3.10.4 and TensorFlow 2.11.0, leveraging the computational power of two Nvidia A100 GPUs for all training and testing processes. This setup ensured efficient model development and evaluation. We call the measure based on the final model AutoCumulus.

### 2.5 Model Testing and Comparisons

We compared the AutoCumulus measures with the original measures by plotting them against each other and calculating the correlation coefficient for a validation subset of 891 images and by plotting the difference between the two measures as a function of their mean (a Bland-Altman plot also known as a Tukey mean-difference plot) and the limits of agreement defined as 2 standard deviations of the difference about the mean difference.

We conducted analyses comparing measures for left and right breast using the independent CSAW-CC dataset risk (Strand, n.d.) which comprises four images (MLO and CC views for both the left and right breasts) for a total of 24,694 women, comprising 1,868 women with and 22,868 women without breast cancer. We exclusively utilized the CC view images. These images had been measured independently for percent density using the LIBRA software.

Using the CSAW-CC dataset, we assessed the relative performances of the AutoCumulus and LIBRA measures by comparing left and right breast images for 1,826 women with and 22,868 women without breast cancer. We compared predictions of interval breast cancer risk using the Area under the Receiver Operating Characteristic curve (AUC) using images for 267 women with and 22,868 women without interval breast cancer.

## 3. Results

Figure 2.A shows the similarity between the estimated automated percent density from the model and the actual percent density measured by humans using a semi-automated method. The Pearson correlation was 0.95.

**Figure 2.**
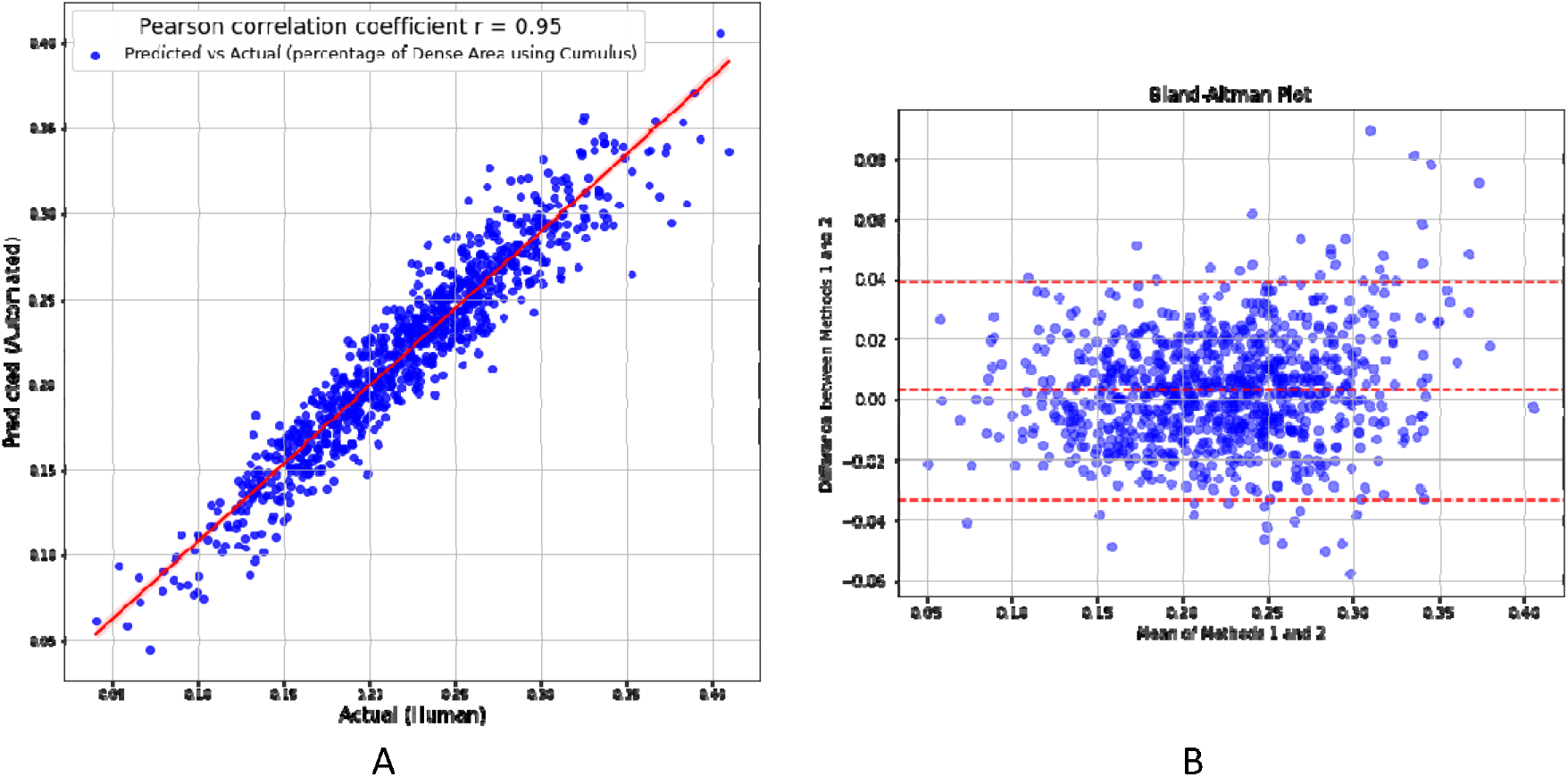
A) Correlation between the automated percent density estimated by AutoCumulus and the corresponding actual measures, B) the Bland-Altman plot, which assess the agreement between the estimated percent density and the corresponding actual measures.

Figure 2.B displays the Bland-Altman plot and shows the measures differed little on average and the limit of agreement was 0.04 and independent of the mean except at the upper tail.

Furthermore, our model underwent testing using the independent CSAW-CC dataset, for which percent density values were pre-measured using the Libra software (Keller et al., 2015). Given the absence of manual measures of percent density in the CSAW-CC dataset, the most effective evaluation method involved assessing the correlation between the percent density values for the left and right breasts (bilateral comparison). This correlation is illustrated in Figure 3.A and 3.B, where percent density was measured using AutoCumulus and LIBRA, respectively. Figure 3.A shows that the correlation between measures for the left and right breast had a correlation of 0.95 when using AutoCumulus, greater than 0.79 when using the LIBRA software (P<0.001).

**Figure 3.**
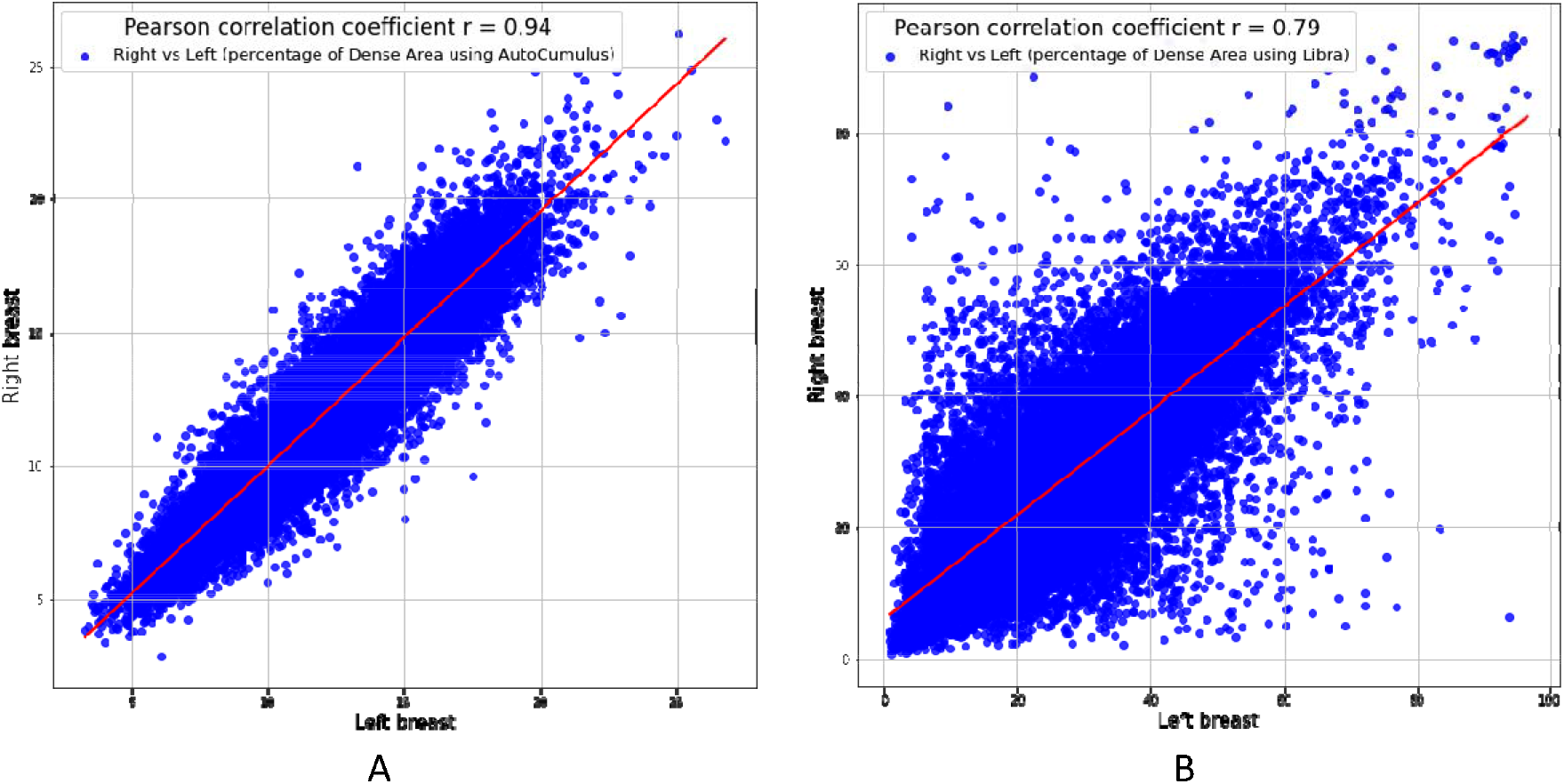
Correlation analysis depicting the relationship between percent density measurements for the left and right breasts using A) AutoCumulus and B) LIBRA.

Figure 4 shows the performance of the average percent density across left and right breasts as a predictor of interval cancer was stronger when using AutoCumulus than LIBRA (AUC = 0.638 (p<0.001) versus 0.597 (p<0.001)). Importantly, performance was better at high levels of specificity (e.g. 95%, equivalent to a low false positive rate of 5%), for which the true positive was 13% for AutoCulumulus and 10% for LIBRA (p<0.001).

**Figure 4.**
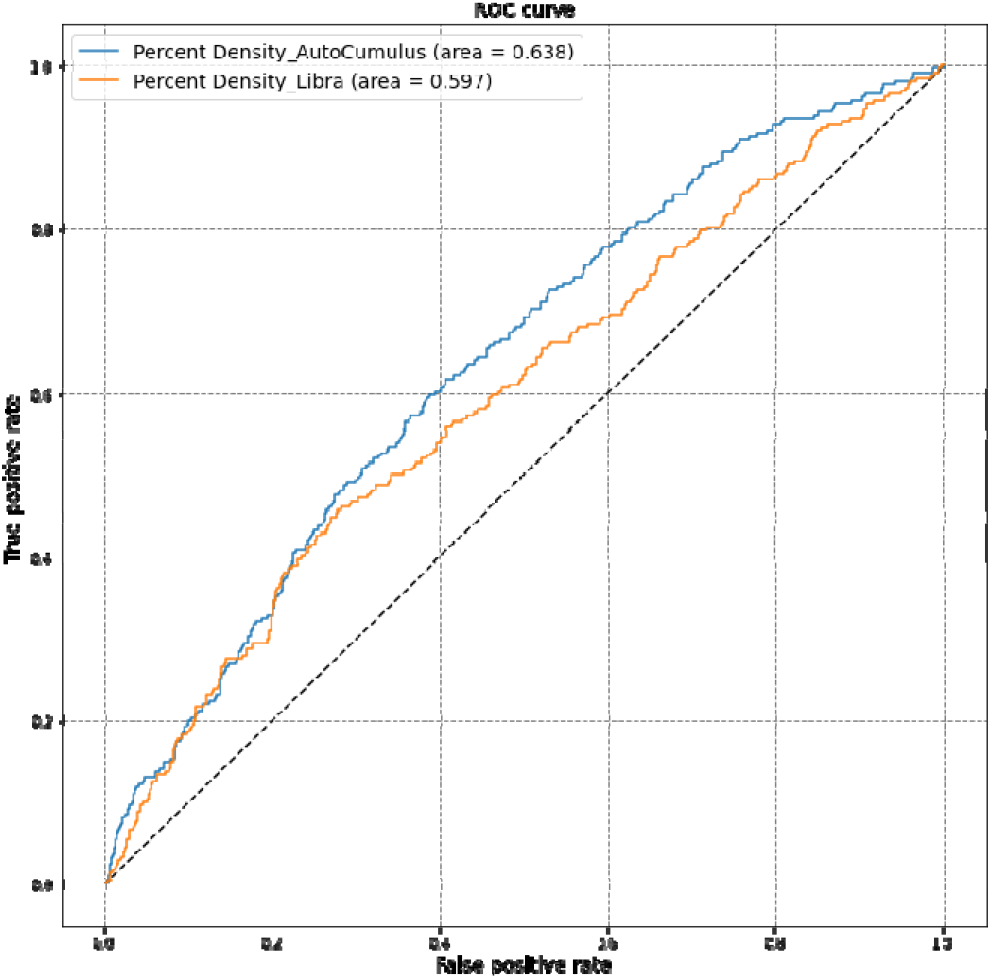
illustrates the (ROC) curves and corresponding (AUC) values for AutoCumulus and Libra as predictors of interval cancer.

## 4. Discussion

We have demonstrated how an automated algorithm for measuring mammographic density can be learnt by applying AI to human-derived measures based on a semi-automated approach using the CUMULUS software. Our approach was validated using an independent dataset form which we found that our automated measure, AutoCumulus, provided more repeatable measures and could predict interval cancers better than the freely available and widely used automated tool LIBRA. One reason behind this could be that AutoCumulus measures were much more strongly correlated between a woman’s left and right breasts.

There is a well-established association between mammographic density and risk of interval cancers that are diagnosed between regular mammographic screens (Kerlikowske et al., 2015; Strand et al., 2019), and there is a wide concern about ‘dense breasts’ per se (REF). Measurement of density has been problematic, however, and clinical practice has generally relied on the pathologists’ interpretation without any formal quality control; the risk predicting performance of the BI-RADS categorisations when used in practice by multiple readers across a period of time is far less than found by controlled research studies utilising one or a few radiologists (Hopper et al., 2020). The specialist semi-automated measure, CUMULUS, is used in many research papers but does not appear to be used in clinical practice. Automated measures such as VOLPARA and LIBRA have been developed and are being used in clinical and research settings.

Strengths of this study include the validation on both internal and external datasets, comparison with an established tool, and the large sample sizes and highly significant findings.

Limitations of this study include use of one experienced measurer, so using a range of measurers could enhance accuracy and broaden applicability. The focus on CC views could limit accuracy so MLO views should also be considered. Other machines should also be studied to broaden clinical reach.

Our findings contribute valuable insights into the potential clinical utility of AutoCumulus in identifying women at risk of interval breast cancer, presenting a promising avenue for future research and clinical applications. They illustrate that the integration of AI into mammographic density assessment represents a significant advancement in overcoming the limitations associated with traditional methods. The transition from reliance on subjective BI-RADS categorizations to more objective and automated measures like AutoCumulus marks a paradigm shift in the field and supports the integration of Deep Learning in the ongoing battle against breast cancer.

## Data Availability

All data produced in the present study are available upon reasonable request to the authors

## Acknowledgement

This work was supported by grants from the Australian Government Medical Research Future Fund (MRFAI000090), the Cancer Council Victoria (AF7305) and the National Breast Cancer Foundation (IIRS-18-093). JLH was supported by an NHMRC Fellowship grant (GMT1137349), SL was supported by a Victoria Cancer Agency Early Career grant (ECRF19020) and an NHMRC Emerging Leadership Fellowship (GNT2017373), and GC was supported by an ARC Future Fellowship grant (FT190100525).

This research was supported by The University of Melbourne’s Research Computing Services and the Petascale Campus Initiative.

